# Machine learning-Based Classification of Papillary Thyroid Carcinoma Versus Multinodular Goiter Using Preoperative Laboratory and Cytology Data

**DOI:** 10.1101/2025.05.15.25327670

**Authors:** Salar GolmohammadzadehKhiaban, Mehrad Namazee, Ali Rahnama

## Abstract

**Background:** Thyroid nodules are frequently encountered in clinical practice, with their detection increasing due to advancements in imaging modalities. While most nodules are benign, distinguishing papillary thyroid carcinoma (PTC) from benign entities such as multinodular goiter (MNG) remains a diagnostic challenge. Fine-needle aspiration (FNA) and sonography are standard tools, but their limitations highlight the need for supplementary approaches. This study evaluates the use of machine learning (ML) models to classify PTC versus MNG using routine preoperative clinical, laboratory, and cytological data before performing surgery and Pathology results.

**Methods:** This retrospective multicenter study included 971 patients who underwent total thyroidectomy between 2020 and 2024. The dataset incorporated demographic data, preoperative sonographic findings, hematologic and thyroid function tests, and FNA cytology results. Five supervised ML algorithms—Logistic Regression, Random Forest, XGBoost, Support Vector Machine (SVM), and K-Nearest Neighbor (KNN)—were trained and validated. Model performance was assessed using accuracy, precision, recall, F1-score, sensitivity, specificity, and area under the receiver operating characteristic curve (AUC-ROC).

**Results:** The XGBoost model achieved the best performance, with an accuracy of 84.4%, precision of 85.3%, and an AUC-ROC of 0.881. It also demonstrated high sensitivity (0.714) and specificity (0.944). Random Forest also performed well (accuracy: 81.2%, AUC-ROC: 0.919). Logistic Regression, SVM, and KNN underperformed in comparison. Feature importance analysis revealed that the FNA result, nodule size, and TSH were the most influential predictors.

**Conclusion:** Machine learning models, particularly XGBoost and Random Forest, show promise in accurately distinguishing between MNG and PTC using routine clinical data. Their integration into preoperative assessment may enhance diagnostic precision, reduce unnecessary procedures, and support personalized surgical decision-making. Further validation in diverse, multicenter cohorts is warranted to confirm generalizability and clinical utility.

## Introduction

Thyroid nodules are highly prevalent clinical findings, and their detection rates increase progressively with the advancement of imaging modalities (1). Although the majority of these nodules are benign (2), a minority may represent malignant lesions, the most common of which is papillary thyroid carcinoma (PTC) (3). Accurate preoperative discrimination between malignant and benign thyroid nodules is essential to avoid unnecessary surgery, reduce patient anxiety, and guide appropriate clinical management (4).

The most prevalent benign thyroid condition, known as multinodular goiter (MNG), is characterized by the thyroid gland containing multiple nodules of various sizes and shapes (5). Despite being the gold standard for the preoperative diagnosis of thyroid nodules, fine-needle aspiration (FNA) cytology can produce non-diagnostic or indeterminate results, especially because of sampling errors and nodule heterogeneity (6). Currently, molecular tests have improved diagnostic accuracy; however, they do not fully eliminate the risk of misdiagnosis or ensure adequate monitoring (7). Despite the ready availability of thyroid function tests and other preoperative laboratory parameters, they are not optimally utilized in diagnostic modeling (8).

In this regard, artificial intelligence (AI) and machine learning have developed as incredibly powerful tools for the enhancement of clinical decision-making (9). These technologies are specifically very good at identifying sophisticated, nonlinear patterns in high-dimensional data, abilities especially valuable in the heterogenous domain of thyroid disease (10, 11). AI models can aggregate and process diverse inputs, such as preoperative laboratory findings and cytological findings, to generate predictive information that may be elusive to conventional statistical methods or clinical judgment (12, 13).

Most existing models rely on radiologic imaging or genomic markers, leaving broadly unaddressed the possibility of cheap, cost-saving AI tools based on data already integrated into standard clinical processes (14, 15). However, there is still a lack of research on the use of machine learning models specifically trained on routine preoperative laboratory and FNA data to differentiate between MNG and PTC. To address this gap, a machine learning model for preoperative stratification of MNG and PTC based on routinely ordered laboratory tests and FNA cytology results was developed and validated. This model seeks to increase risk stratification, improve diagnostic accuracy, and facilitate better surgical decision-making by leveraging AI.

## Method

### 1. Study Design and Population

Patients who underwent thyroid surgery between 2020 and 2024 at three major tertiary care centers were included. Inclusion criteria were as follows: over 18 years patients who underwent thyroid surgery with a confirmed histopathological diagnosis, had available preoperative laboratory test results, and had undergone fine-needle aspiration (FNA) cytology. Patients with incomplete preoperative data, a history of prior thyroid surgery, or final histopathology revealing non-papillary histology’s were excluded. Finally, a total of **971 patients** met the eligibility criteria, including 408 cases of MNG and 553 cases of PTC

### 2. Data Collection and Preprocessing

Data were retrospectively collected from the electronic medical records of Firoozgar and Rasoul Akram Hospitals between 2020 and 2024. Only patients who underwent total thyroidectomy and had complete records for the required variables were included. Cases with missing key features were excluded from the analysis.

The dataset included demographic information (age, gender), Sonographic findings (nodule size and location, lymph node status), preoperative laboratory values (complete blood count, thyroid function tests, calcium), cytology results from preoperative fine-needle aspiration (FNA) including Benign, Malignant, papillary thyroid carcinoma (PTC), AUS, Not-deterministic and also final surgical pathology (benign vs. malignant, tumor type).

Prior to model development, all numeric variables were standardized using z-score normalization. Categorical variables were encoded using one-hot encoding where applicable. Outliers and biologically implausible values were reviewed and removed. To ensure data consistency, all preprocessing steps were performed using reproducible Python-based pipelines.

### 3. AI Model Development and Evaluation

The objective of model development was to classify patients with papillary thyroid carcinoma versus multinodular goiter using preoperative clinical, laboratory, cytological, and imaging data. Multiple supervised learning algorithms were explored to identify the most effective classifier for this task.

The dataset was randomly split into training (80%) and testing (20%) sets while preserving class balance. Feature selection was performed using both domain knowledge and data-driven techniques, including univariate statistical tests and feature importance scores derived from tree-based models. To address class imbalance and improve generalizability, stratified sampling was used during cross-validation.

Several machine learning algorithms were implemented and compared, including:

- Logistic Regression
- Random Forest
- Gradient Boosting Machines (XGBoost)
- Support Vector Machine (SVM)
- K-Nearest Neighbor (KNN)

Model development and tuning were carried out using scikit-learn (16), XGBoost (17) and data management, visualization using NumPy(18) and Matplotlib(19) in Python. Hyperparameters were optimized via grid search with 5-fold cross-validation on the training set. Performance metrics such as accuracy, precision, recall, F1-score, Sensitivity-Specificity and area under the receiver operating characteristic curve (AUC-ROC) were used to evaluate each model on the independent test set.

### 4. Ethical Consideration

This study was conducted in accordance with the Declaration of Helsinki and approved by the Ethics Committees of Firoozgar Hospital and Rasoul Akram Hospital [the institutional review board (IRB) Number of 20-169-2024] The study utilized de-identified retrospective data, and informed consent was waived due to the non-interventional nature of the research. All data handling procedures adhered to institutional and national regulations on data privacy and patient confidentiality.

## Result

### 1. Patient Characteristics

The study cohort included 951 patients (~59% Women / ~41% Men) with a mean age of 45.85 years (range: 15–78). The average nodule size was 33.02 mm, with a wide variation from 5 mm to 96 mm. Hematologic parameters such as white blood cell count (WBC), hemoglobin (Hb), and platelet count (PLT) had means of 7.97 × 10^9^/L, 13.98 g/dL, and 235.09 × 10^9^/L, respectively. Thyroid function tests showed an average TSH level of 2.42 μIU/mL and total T4 (tt4) of 9.84 μg/dL. Full summary statistics for clinical and laboratory variables are presented in **Table 1**.

**Table 1.** Descriptive statistics of patient characteristics. Shown are the minimum, maximum, mean, median, and standard deviation for demographic, hematologic, and endocrine variables.

| Variable | Min | Max | Mean | Median | Std Dev |
| --- | --- | --- | --- | --- | --- |
| Age (years) | 15.00 | 78.00 | 45.85 | 45.50 | 13.39 |
| Size (mm) | 5.00 | 96.00 | 33.02 | 30.00 | 18.75 |
| WBC ($\times 10^9/L$) | 3.80 | 19.70 | 7.97 | 7.50 | 2.72 |
| NP (%) | 9.10 | 96.30 | 66.05 | 64.50 | 13.51 |
| NC ($\times 10^9/L$) | 1.40 | 66.40 | 5.72 | 4.60 | 5.48 |
| LP (%) | 2.30 | 61.00 | 26.67 | 28.85 | 11.33 |
| Lymph Count | 0.10 | 29.40 | 2.21 | 1.90 | 2.47 |
| NTL Ratio | 0.53 | 42.00 | 4.13 | 2.17 | 5.58 |
| Hemoglobin (g/dL) | 9.10 | 119.00 | 13.98 | 13.20 | 8.64 |
| PLT ($\times 10^9/L$) | 105.00 | 686.00 | 235.09 | 225.50 | 65.42 |
| RDW (%) | 11.00 | 20.00 | 13.68 | 13.10 | 1.74 |
| PDW (%) | 8.20 | 19.70 | 12.07 | 11.60 | 2.14 |
| MPV (fL) | 1.40 | 12.20 | 9.41 | 9.40 | 1.35 |
| TSH ($\mu IU/mL$) | 0.01 | 29.30 | 2.42 | 1.40 | 3.89 |
| TT4 ($\mu g/dL$) | 0.10 | 139.00 | 9.84 | 7.90 | 14.16 |
| TT3 (ng/dL) | 0.76 | 242.00 | 93.28 | 105.00 | 59.19 |
| Calcium (mg/dL) | 7.10 | 12.00 | 9.26 | 9.30 | 0.83 |

### 2. Models Performance

In this study, five machine learning models were trained and evaluated to classify thyroid pathology using preoperative laboratory features. The models included Logistic Regression, Random Forest, XGBoost, Support Vector Machine (SVM), and K-Nearest Neighbors (KNN). The performance of each model was assessed using several metrics as summarized in **Table 2**. Additionally, sensitivity and specificity were computed to evaluate the models’ clinical discriminative abilities (Table 2).

**Table 2.** Performance metrics of five machine learning models for thyroid nodule classification. The table summarizes accuracy, precision, recall, F1-score, AUC-ROC, sensitivity, and specificity. XGBoost and Random Forest achieved the highest overall performance across most metrics.

| Model | Accuracy | Precision | Recall | F1-score | AUC-ROC | Sensitivity | Specificity |
| --- | --- | --- | --- | --- | --- | --- | --- |
| XGBoost | 0.844 | 0.853 | 0.844 | 0.840 | 0.881 | 0.714 | 0.944 |
| Random Forest | 0.812 | 0.828 | 0.812 | 0.806 | 0.919 | 0.643 | 0.944 |
| Logistic Regression | 0.719 | 0.720 | 0.719 | 0.713 | 0.671 | 0.571 | 0.833 |
| SVM | 0.688 | 0.703 | 0.688 | 0.667 | 0.639 | 0.429 | 0.889 |
| KNN | 0.562 | 0.556 | 0.562 | 0.557 | 0.625 | 0.429 | 0.667 |

XGBoost achieved the highest overall performance, with an accuracy of 84.4%, precision of 85.3%, recall of 84.4%, F1-score of 84.0%, and an AUC-ROC of 0.881. Importantly, it also demonstrated high sensitivity (0.714) and specificity (0.944), indicating a strong ability to correctly identify both positive and negative cases. Random Forest performed comparably well, with an accuracy of 81.2% and the highest AUC-ROC of 0.919 among all models. It also exhibited strong specificity (0.944), although its sensitivity (0.643) was slightly lower than that of XGBoost. (Table 1)

Logistic Regression showed moderate performance (accuracy: 71.9%, AUC-ROC: 0.671) and a notable imbalance between sensitivity (0.571) and specificity (0.833), suggesting it was more conservative in predicting positive cases.

SVM and KNN underperformed relative to the other classifiers. SVM achieved an accuracy of 68.8% and a low AUC-ROC of 0.639 showed the lowest performance across all metrics, with an accuracy of 56.2%, AUC-ROC of 0.625, and similarly poor sensitivity A visual comparison of the classifiers’ performance is presented in **Figure 1**, which shows the ROC curves for all five models. Ensemble methods such as XGBoost and Random Forest demonstrated steeper curves and larger AUC values, reflecting their superior classification performance

**Figure 1.**
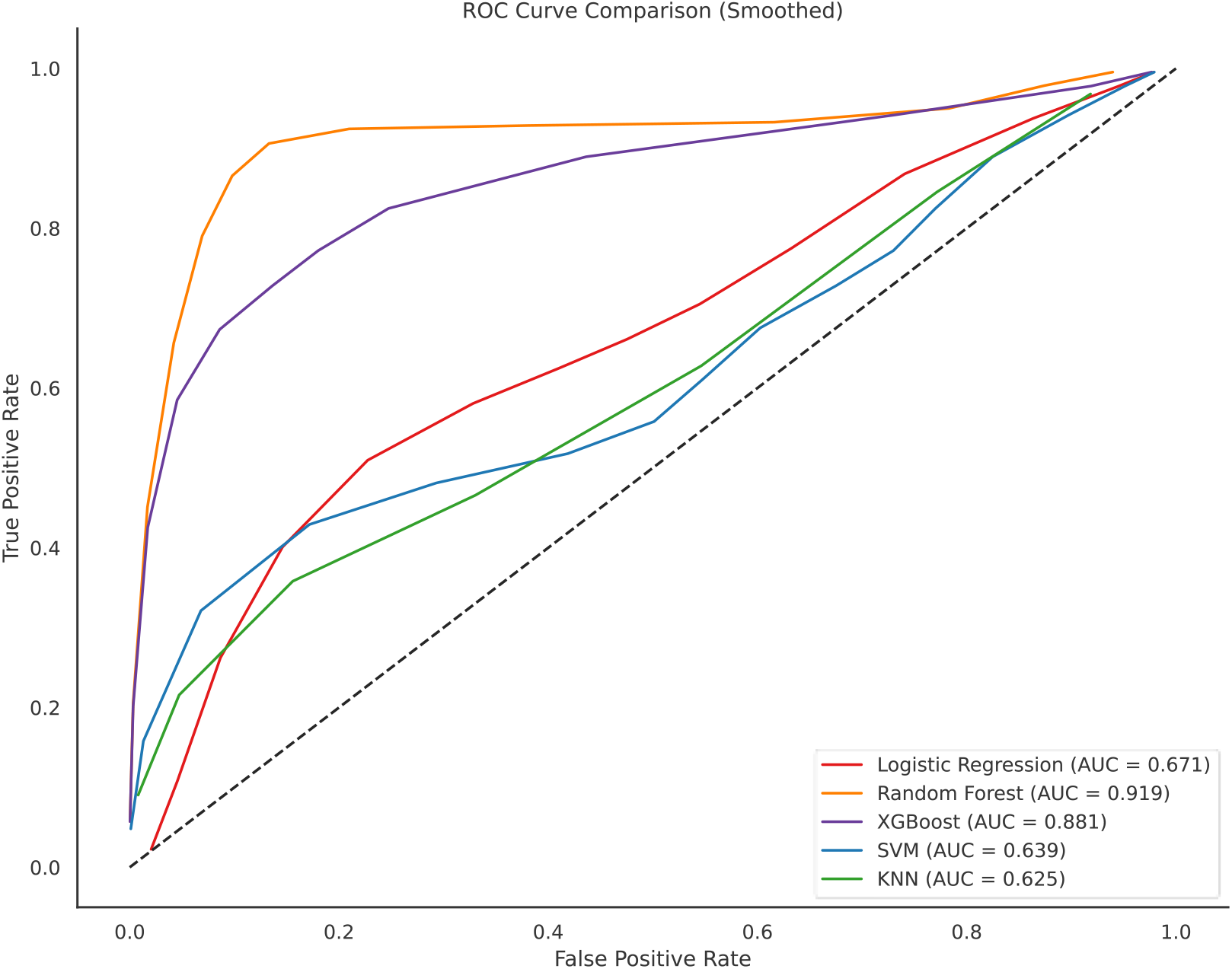
ROC curves of five machine learning models used for classifying thyroid nodules. XGBoost and Random Forest models demonstrated the AUC values (0.881 and 0.919, respectively), indicating superior discriminatory performance compared to other models. curves smoothed using a gaussian kernel with sigma=1 for better visualization.

### 3. Feature Importance

To gain insight into the contribution of each feature to the model’s predictive performance, we evaluated feature importance using the trained XGBoost classifier. The results are presented in **Figure 2**.

**Figure 2.**
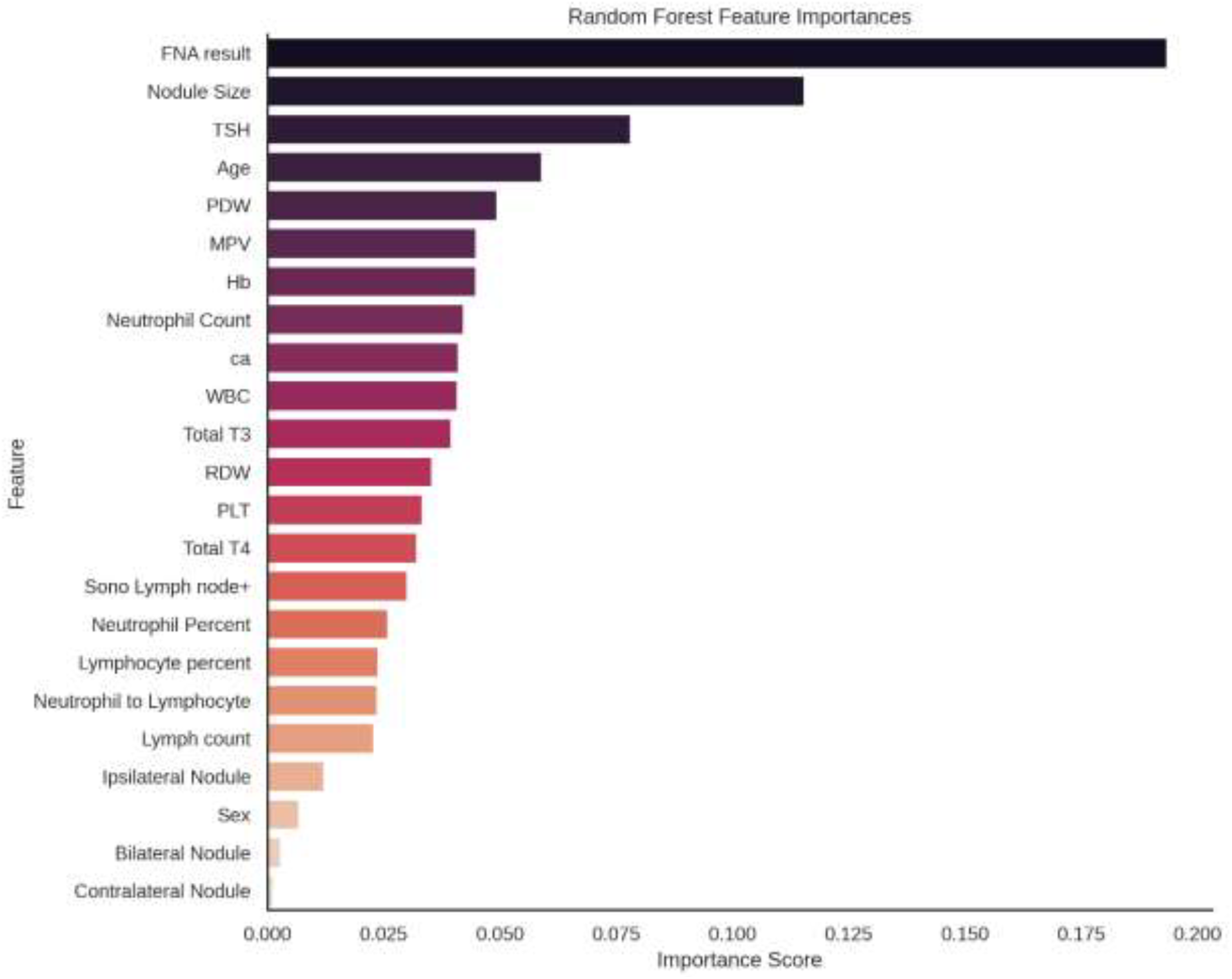
Feature importance scores derived from the XGBoost model. The plot ranks input variables based on their relative contribution to model predictions. FNA result, nodule size, and TSH were among the most influential features, while demographic and anatomical variables such as sex and contralateral nodule presence showed minimal importance.

Among all input variables, the fine needle aspiration (FNA) result was the most influential predictor of thyroid pathology, with an importance score of 0.194, followed by nodule size (0.116), and thyroid-stimulating hormone (TSH) levels (0.078). These were followed by age, platelet distribution width (PDW), mean platelet volume (MPV), and hemoglobin (Hb), each contributing moderate importance to the classification task.

In contrast, features such as **sex, bilateral nodule**, and **contralateral nodule presence** had minimal influence on model performance, each with importance values below **0.01**. These findings emphasize the dominant role of cytological and imaging characteristics (e.g., FNA result, nodule size), as well as selected laboratory biomarkers, in distinguishing malignant from benign thyroid nodules in our dataset.

## Discussion

Thyroid nodules are one of the most common clinical findings, with their prevalence increasing due to advancements in imaging techniques (20). While the majority of thyroid nodules are benign, distinguishing malignant nodules, particularly PTC, remains a critical clinical challenge(21). This study presents a machine learning (ML) model that integrates routine preoperative clinical, laboratory, and FNA cytology data to classify thyroid nodules as either benign (MNG) or malignant (PTC), which holds significant potential for improving diagnostic accuracy and patient outcomes.

The diagnosis and management of thyroid nodules heavily rely on imaging modalities such as ultrasound and, when necessary, FNA cytology. However, these methods have limitations, including operator dependency and the risk of non-diagnostic or indeterminate results. Molecular tests, though improving diagnostic accuracy, are often costly and do not fully eliminate the risk of misdiagnosis (21). Additionally, while thyroid function tests and other laboratory parameters are readily available, they have not been optimally integrated into predictive models for preoperative stratification of thyroid nodules. This study addresses this gap by incorporating preoperative laboratory test results, sonographic findings, and cytological data into an ML framework. Such integration is crucial because it allows for the inclusion of a diverse set of inputs, reflecting the multifactorial nature of thyroid disease. ML algorithms, particularly those suited for high-dimensional data, excel at recognizing complex, nonlinear relationships among variables, providing a more comprehensive and potentially more accurate diagnostic tool than traditional methods. The model’s use of routinely collected clinical data enhances its accessibility and cost-effectiveness, making it a practical alternative to more resource-intensive diagnostic approaches (22).

The performance of the ML models developed in this study was evaluated across several metrics, including accuracy, precision, recall, F1-score, sensitivity, specificity, and AUC-ROC. The XGBoost classifier performed the best among the five models tested, achieving an accuracy of 84.4%, precision of 85.3%, and an AUC-ROC of 0.881. These results indicate that the model is highly effective at distinguishing between MNG and PTC. Furthermore, the high sensitivity (0.714) and specificity (0.944) suggest that the model is capable of both correctly identifying malignant cases (sensitivity) and correctly ruling out benign ones (specificity).

In comparison, Random Forest also performed well, with an accuracy of 81.2% and the highest AUC-ROC of 0.919, though its sensitivity was slightly lower than that of XGBoost. Logistic Regression, Support Vector Machine (SVM), and K-Nearest Neighbor (KNN) underperformed relative to these two models, with Logistic Regression showing a notable imbalance between sensitivity and specificity. The relatively lower performance of SVM and KNN indicates that more complex models like XGBoost and Random Forest are better suited for this classification task, likely due to their ability to handle nonlinear relationships and interactions among features. These findings align with previous studies where ML models, particularly ensemble methods like Random Forest and gradient boosting, have demonstrated robust performance in classifying thyroid nodules based on clinical and imaging data(23). For instance, a study by Liu et al. (2022) developed an AI model with an AUC-ROC of 0.91 for distinguishing malignant thyroid nodules, further supporting the reliability of ML in this domain (24). This consistency across studies suggests that machine learning has significant potential in enhancing the accuracy and efficiency of thyroid nodule diagnosis.

A key aspect of this study is the identification of the most important features contributing to the model’s performance. Feature importance analysis revealed that the FNA result, nodule size, and TSH levels were the most influential predictors of thyroid pathology. These findings are consistent with clinical knowledge, where cytological features (such as FNA results) and nodule characteristics (such as size) are crucial for evaluating the risk of malignancy. TSH levels, a well-established biomarker for thyroid function, further support the biological relevance of these features in distinguishing benign from malignant nodules (25). In contrast, demographic factors such as sex and the presence of contralateral nodules had minimal impact on model performance. This is important because it demonstrates that the model successfully prioritizes clinically meaningful features while minimizing the influence of less relevant variables. These results suggest that the model’s decision-making process closely mirrors the clinical decision-making process, where FNA and nodule characteristics are paramount, and demographic factors play a secondary role.

The clinical implications of this study are profound. First, the ability to preoperatively distinguish between MNG and PTC can significantly influence treatment decisions. By identifying patients at higher risk for malignancy, clinicians can make more informed decisions regarding the need for surgery, the extent of the procedure, and postoperative care. For instance, benign nodules may warrant conservative monitoring, while malignant nodules may require more aggressive interventions, including total thyroidectomy and lymph node dissection. Furthermore, reducing unnecessary surgeries and biopsies can alleviate patient anxiety and reduce healthcare costs (23). As the model utilizes routine clinical data that are already collected as part of standard practice, its implementation would not require significant changes to current clinical workflows, making it a feasible tool for widespread adoption. This approach could also enhance patient care by providing more personalized management based on individual risk profiles, thus improving overall healthcare efficiency.

While the model demonstrates promising results, there are several limitations to consider. The retrospective design and reliance on data from a single institution may limit the generalizability of the findings. To enhance the robustness of the model, future studies should validate the model in diverse, multicenter cohorts to ensure its applicability across different populations and settings. Moreover, the model could be further improved by incorporating additional data, such as genomic markers, advanced imaging features, and patient-reported outcomes, which may provide a more comprehensive understanding of thyroid nodule pathology. Future research could also explore the integration of AI-generated content-enhanced computer-aided diagnosis (AIGC-CAD) systems, which would allow for more advanced diagnostic workflows and facilitate human-machine collaboration in clinical decision-making.

## Conclusion

This study highlights the feasibility and efficacy of using machine learning models based on routine preoperative clinical and laboratory data to differentiate between benign and malignant thyroid nodules. The integration of such models into clinical practice could revolutionize preoperative stratification, leading to more personalized and efficient patient care. As healthcare systems increasingly embrace digital tools, AI-driven models hold the potential to significantly improve diagnostic accuracy, reduce unnecessary procedures, and ultimately enhance patient outcomes. Further research and validation are essential to refine these models and ensure their widespread applicability in diverse healthcare settings.

## Data Availability

All data produced in the present study are available upon reasonable request to the authors

